# Blood transcriptomics reveals a Parkinson’s disease signature and heterogeneous prodromal molecular profiles in isolated RBD

**DOI:** 10.64898/2026.06.30.26356917

**Authors:** Peter Artimovic, Kristina Kulcsarova, Marek Kloc, Monika Svecová, Eva Feketeova, Milan Maretta, Petronela Christova, Barbora Zecova, Ema Kerpcarova, Miriama Ostrozovicova, Stefan Orkuty, Jana Papikova, Matej Skorvanek, Miroslava Rabajdova

## Abstract

**Background:** Parkinson’s disease (PD) has a prolonged prodromal phase, but minimally invasive molecular biomarkers distinguishing manifest PD from prodromal synucleinopathy remain insufficiently characterized. Isolated REM sleep behavior disorder (iRBD) represents a high-risk prodromal condition and provides an opportunity to investigate early blood-based transcriptional alterations.

**Objective:** To identify peripheral blood transcriptomic signatures distinguishing healthy controls (HC), individuals with iRBD, and patients with PD, and to explore whether longitudinal iRBD samples exhibit movement toward a PD-like transcriptional state.

**Methods:** Peripheral blood RNA-seq data were analyzed using harmonized metadata, DESeq2 differential-expression analysis, internally validated machine-learning models, PD-like projection, and integrated biomarker-panel prioritization. Independent baseline samples were used for cross-sectional differential-expression and machine-learning analyses. iRBD follow-up and post-conversion observations were excluded from baseline model development and reserved for exploratory longitudinal analyses.

**Results:** Baseline analyses included 71 independent samples: 20 HC, 31 iRBD, and 20 PD. An additional 19 iRBD follow-up observations, including three post-conversion observations, were available for exploratory analyses. Differential-expression analysis identified 170 FDR-significant genes in PD versus HC and 85 in PD versus iRBD, compared with one FDR-significant gene in iRBD versus HC. Internal machine-learning validation showed stronger discrimination of manifest PD, with a best ROC-AUC of 0.883 for HC versus PD and 0.889 for iRBD versus PD. Discrimination between HC and iRBD was weak, with a best ROC-AUC of 0.584. PD-like projection scores were lowest in HC, highest in PD, and heterogeneous among baseline iRBD samples. Follow-up iRBD samples showed an exploratory upward shift in the mean PD-like projection score. Integrated prioritization produced a 24-gene PD candidate panel and a 24-gene exploratory iRBD panel, with genes in each panel supported by machine-learning feature-stability evidence and differential expression analysis.

**Conclusions:** Manifest PD was associated with a distinct peripheral blood transcriptional signature, whereas iRBD-associated alterations were substantially weaker and more heterogeneous. The prioritized panels represent candidates for independent technical and external validation and should not yet be interpreted as clinically validated diagnostic or prognostic tests.

## Introduction

Parkinson’s disease (PD) is a progressive and clinically heterogeneous neurodegenerative disorder and a major neuronal α-synucleinopathy. Although PD is currently still diagnosed only after the emergence of characteristic motor signs, the underlying neurodegenerative process begins years or even decades earlier. Contemporary models conceptualize PD as a biological continuum comprising an asymptomatic preclinical phase followed by a prodromal stage characterized by the gradual emergence of non-motor and subtle motor symptoms before the development of clinically manifest parkinsonism. These prodromal manifestations may include REM sleep behaviour disorder (RBD), hyposmia, autonomic dysfunction, neuropsychiatric symptoms, and subtle cognitive or motor changes (1,2). Importantly, PD is increasingly recognized as a biologically heterogeneous disorder rather than a single uniform disease entity. Emerging models suggest the existence of distinct disease trajectories, including proposed body-first and brain-first subtypes, which may differ in the anatomical origin and subsequent spread of α-synuclein (α-syn) pathology, resulting in distinct sequences of pathological events, clinical manifestations, and biomarker profiles. These differences are likely most evident during the prodromal phase, when the distribution of pathology remains relatively restricted and produces markedly different clinical and biological phenotypes (3–5).

This extended and biologically heterogeneous disease continuum creates an opportunity for earlier disease detection, biological stratification, and ultimately preventive or therapeutic disease-modifying interventions. At the same time, it highlights the need for accessible biomarkers capable of detecting disease-associated processes before or around the transition to clinically manifest PD. Recent advances in biological disease definition, including the SynNeurGe criteria and the Neuronal α-Synuclein Disease Integrated Staging System (NSD-ISS), have introduced frameworks that allow PD-related pathology to be identified based on biomarkers alone, even before the onset of classical motor symptoms (6,7). However, currently incorporated biological markers remain largely confined to research settings. Although α-syn seed amplification assays (α-syn SAA) have demonstrated remarkable diagnostic performance, their implementation in population-scale screening remains limited by the technical complexity of the assay, the need for further standardization across laboratories, and, in many current applications, the requirement for cerebrospinal fluid or other invasive tissue sampling. In addition, the long-term prognostic significance of α-syn positivity in asymptomatic individuals remains incompletely understood and requires further longitudinal investigation (8).

Consequently, clinical prodromal markers such as isolated RBD (iRBD), hyposmia, and autonomic dysfunction continue to represent the most accessible approach for identifying individuals at increased risk of future PD. Among these, iRBD occurring before an established neurodegenerative diagnosis represents the strongest clinical marker of prodromal synucleinopathy, with longitudinal studies demonstrating over 90% cumulative probability of phenoconversion to PD, dementia with Lewy bodies (DLB), or, less frequently, multiple system atrophy (MSA) (4,9). Although not all patients with PD develop RBD and only a subset experience it before the onset of motor symptoms (5), making iRBD a non-universal prodromal phenotype of PD, likely reflecting specific biological trajectories rather than the entire spectrum of the disease, it provides a unique and clinically accessible opportunity to study disease-associated processes years before the onset of clinically manifest neurodegeneration.

Beyond disease detection, the emergence of biological diagnostic criteria highlights a broader challenge: while α-syn-based biomarkers may enable increasingly accurate identification of synucleinopathy, they provide limited insight into the molecular mechanisms underlying disease initiation, progression, and clinical heterogeneity. The recently proposed biological frameworks explicitly anticipate the future integration of complementary markers reflecting processes such as immune dysfunction, inflammation, mitochondrial and lysosomal impairment, or metabolic dysregulation (6). Understanding these pathways is essential not only for improving biological stratification, but also for identifying mechanisms relevant to future disease-modifying interventions.

From this perspective, peripheral blood transcriptomics provides a minimally invasive approach for investigating systemic molecular changes associated with PD and may serve less as a diagnostic tool and more as a window into disease-associated biological processes operating across different stages of PD development. Earlier studies demonstrated that disease-associated gene-expression signatures can be detected in blood (10,11), and a large longitudinal RNA-sequencing study subsequently identified extensive transcriptional alterations in PD, including persistent changes in neutrophil- and lymphocyte-associated expression programs (12). These observations are consistent with broader evidence implicating peripheral immune dysfunction, inflammatory signaling, metabolic remodeling, and altered circulating immune-cell states in PD (13). Although blood transcriptional profiles should not be considered direct surrogates of brain pathology, they may provide clinically accessible readouts of peripheral disease-associated biology.

Blood-based biomarker discovery nevertheless presents substantial analytical challenges. Whole-blood expression profiles are influenced by leukocyte composition, age, sex, medication exposure, inflammatory comorbidities, sample handling, and technical processing (12,14). Unaccounted technical variation can generate spurious expression differences or obscure genuine biological signals (15). Longitudinal samples introduce additional dependence because repeated observations from the same participant cannot be treated as independent disease cases. In high-dimensional machine-learning analyses, performing feature selection, scaling, or other supervised preprocessing before cross-validation can further produce optimistically biased performance estimates (16). Clear separation of cross-sectional and longitudinal analysis sets, together with leakage-aware model validation, is therefore essential for interpretable biomarker discovery.

Here, we investigated peripheral blood RNA-seq signatures across healthy controls, individuals with iRBD, and patients with manifest PD. Independent baseline samples were used for cross-sectional differential-expression analysis, machine-learning model development, and candidate biomarker prioritization, whereas iRBD follow-up and post-conversion observations were reserved for exploratory longitudinal analyses. We aimed to identify blood transcriptional signatures associated with manifest PD, determine whether a detectable peripheral expression signature is already present in iRBD, and explore how longitudinal transcriptomic profiles evolve during the prodromal phase and following phenoconversion, including whether they shift along a composite PD-like transcriptional axis. The study was designed as candidate biomarker discovery and prioritization for future external validation rather than as the development of a clinically validated diagnostic or prognostic test.

## Results

### Cohort structure and analytical design for blood biomarker discovery

The cross-sectional baseline analysis included 71 independent participants: 20 healthy controls (HC), 31 participants with isolated REM sleep behaviour disorder (iRBD), and 20 patients with Parkinson’s disease (PD). In addition, 19 iRBD follow-up observations, including three post-conversion observations, were available for exploratory longitudinal and projection analyses. Datasets from two batches were harmonized before further analysis (Figure 1).

**Table 1.** Sample inclusion for peripheral blood biomarker discovery analyses.

| Analysis set | N | HC | iRBD | PD | iRBD-FU | Converters |
| --- | --- | --- | --- | --- | --- | --- |
| All samples | 90 | 20 | 50 | 20 | 19 | 3 |
| Independent baseline DESeq2 | 71 | 20 | 31 | 20 | 0 | 0 |
| Baseline ML | 71 | 20 | 31 | 20 | 0 | 0 |
| RBD follow-up metadata | 19 | 0 | 19 | 0 | 19 | 3 |
| PD-like projection | 90 | 20 | 50 | 20 | 19 | 3 |

**Figure 1.**
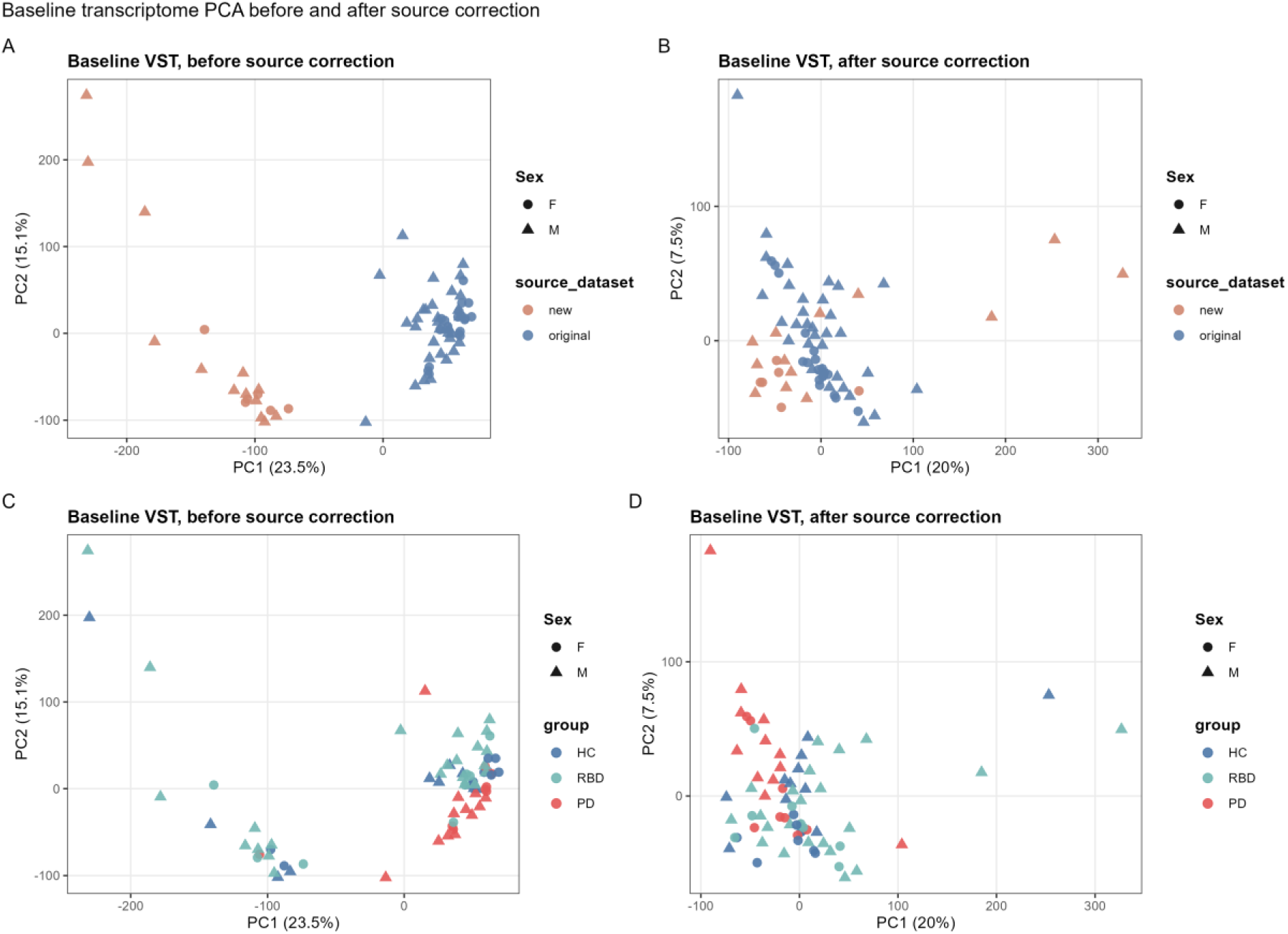
Source-aware batch PCA QC supporting downstream blood biomarker analysis.

### Source-adjusted differential expression identifies a robust PD blood biomarker signal

Using the DESeq2 design ∼ source dataset + group, PD vs HC identified 170 FDR-significant and 1488 nominal genes, and PD vs iRBD identified 85 FDR-significant and 1280 nominal genes. In contrast, iRBD vs HC identified only 1 FDR-significant and 488 nominal genes.

These results indicate that the dominant baseline blood biomarker signal is associated with manifest PD rather than uniform prodromal iRBD status. The contrast pattern indicates that the strongest reproducible peripheral blood signal in this cohort is associated with manifest PD rather than with baseline iRBD. PD remained transcriptionally distinct from both HC and iRBD after adjustment for source dataset, suggesting that the PD-associated signal was not limited to a single reference comparison. In contrast, the limited FDR-level separation between iRBD and HC argues against a uniform cross-sectional iRBD blood signature in the current baseline dataset. This does not exclude prodromal biology in iRBD, but suggests that such signals may be weaker, more heterogeneous across individuals, more dependent on longitudinal change, or more difficult to detect in bulk blood RNA-seq at baseline.

### Internal machine learning supports PD discrimination but not reliable iRBD classification

Random forest provided the best HC-versus-PD discrimination, with a ROC-AUC of 0.883 (95% CI 0.752–0.987), PR-AUC of 0.917, balanced accuracy of 0.825, sensitivity of 0.800, and specificity of 0.850. The model correctly classified 17 of 20 HC and 16 of 20 PD samples. HC-versus-iRBD discrimination was substantially weaker. Logistic regression achieved an ROC-AUC of 0.584, balanced accuracy of 0.617, sensitivity of 0.484, and specificity of 0.750. For iRBD versus PD, logistic regression achieved an ROC-AUC of 0.889, PR-AUC of 0.843, balanced accuracy of 0.764, sensitivity of 0.850, and specificity of 0.677. Results are further summarized in Table 2. Discrimination ability of best models was also represented by the confusion matrices shown in Figure 2.

**Figure 2.**
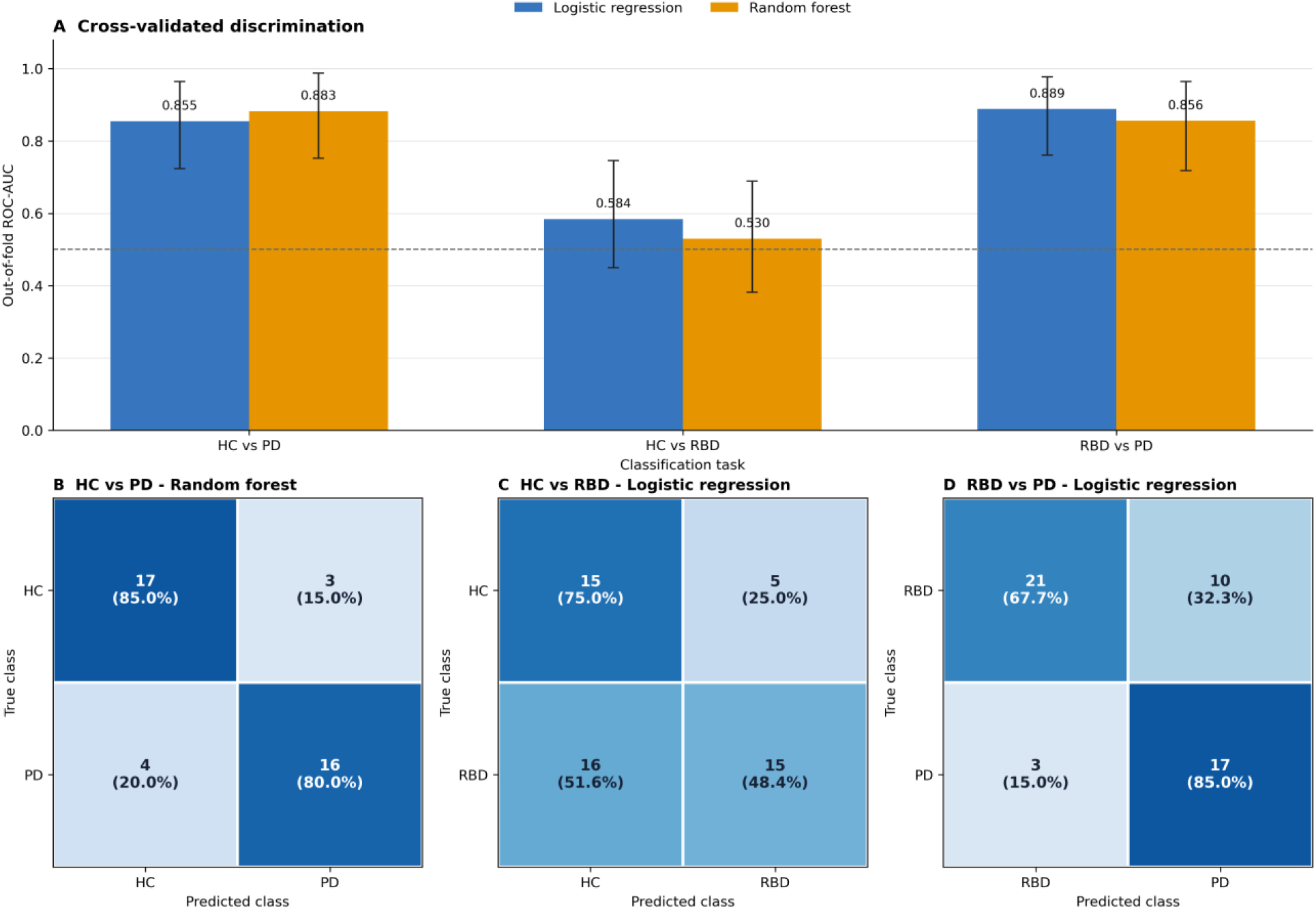
Internal machine-learning performance across diagnostic comparisons. (A) Out-of-fold ROC-AUC values with 95% bootstrap confidence intervals for logistic-regression and random-forest classifiers. The dashed line denotes chance-level discrimination. (B–D) Out-of-fold confusion matrices for the model with the highest ROC-AUC in each comparison. Cells show absolute sample counts and percentages within each true diagnostic class. Threshold-dependent predictions were calculated using a probability threshold of 0.50.

**Figure 3.**
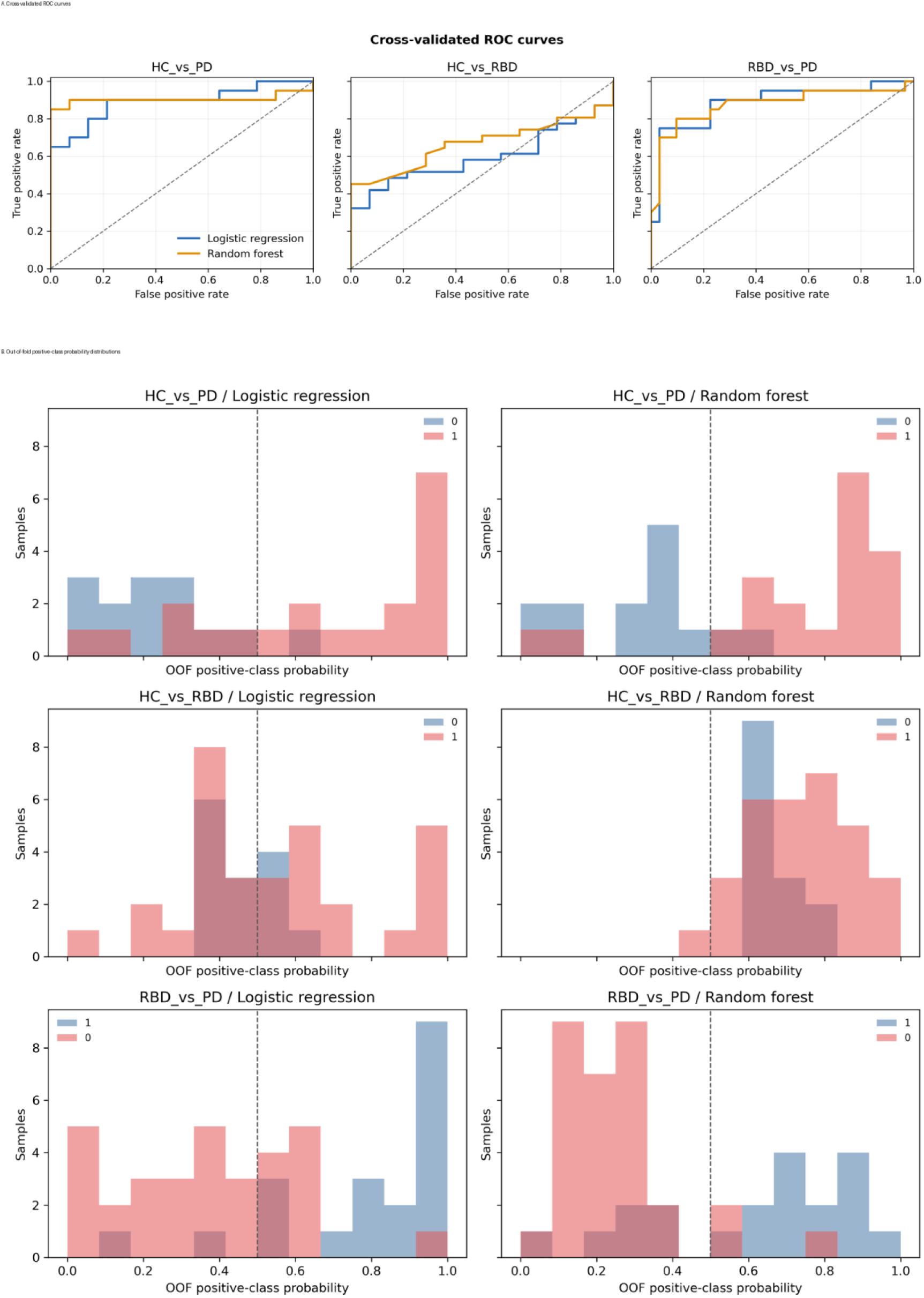
ROC curves and out-of-fold probability distributions for baseline biomarker classifiers.

**Table 2.**
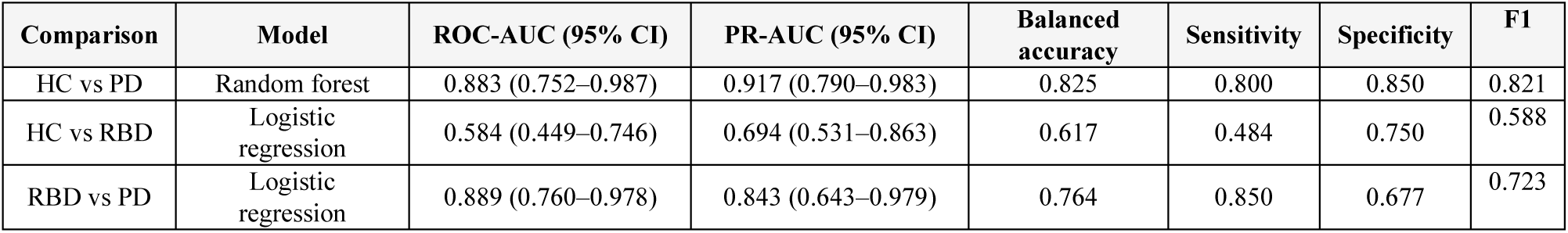
Internally cross-validated ML performance of best models for candidate blood biomarker discrimination.

The ML results therefore paralleled the differential-expression findings. Classifiers separating PD from either HC or iRBD achieved substantially higher internal performance than the HC vs iRBD task, indicating that multigene expression patterns also captured a stronger PD-associated signal than an iRBD-associated baseline signal. In contrast, the modest HC vs iRBD performance, together with wide confidence intervals and weak FDR-level differential expression, argues against interpreting baseline iRBD as a reliably separable diagnostic class in this dataset. These models should therefore be viewed as internally cross-validated support for a PD-associated biomarker pattern, not as externally validated diagnostic classifiers.

### PD-like projection reveals heterogeneous longitudinal blood transcriptomic shifts in iRBD

The longitudinal biomarker question was whether iRBD samples move along a PD-like blood transcriptional axis. HC baseline samples had low projection scores, PD baseline samples had high scores, and baseline iRBD samples were mostly HC-like but heterogeneous. iRBD follow-up observations shifted upward on average, while post-conversion samples were too few for prognostic inference (Figure 4).

**Table 3.**
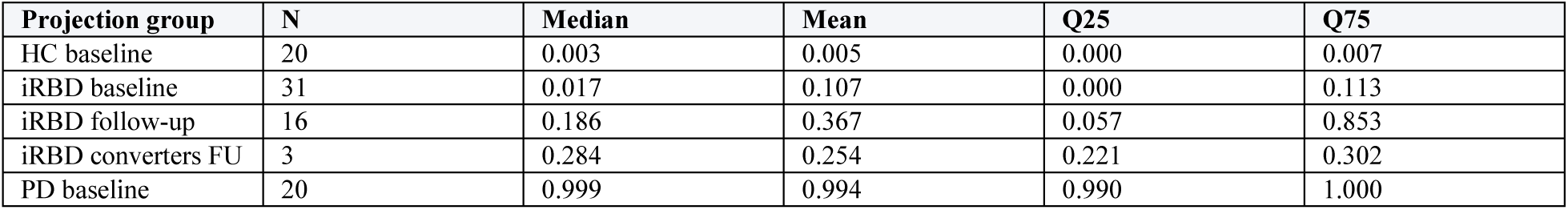
Exploratory PD-like blood transcriptomic projection by group.

| Projection group | N | Median | Mean | Q25 | Q75 |
| --- | --- | --- | --- | --- | --- |
| HC baseline | 20 | 0.003 | 0.005 | 0.000 | 0.007 |
| iRBD baseline | 31 | 0.017 | 0.107 | 0.000 | 0.113 |
| iRBD follow-up | 16 | 0.186 | 0.367 | 0.057 | 0.853 |
| iRBD converters FU | 3 | 0.284 | 0.254 | 0.221 | 0.302 |
| PD baseline | 20 | 0.999 | 0.994 | 0.990 | 1.000 |

**Figure 4.**
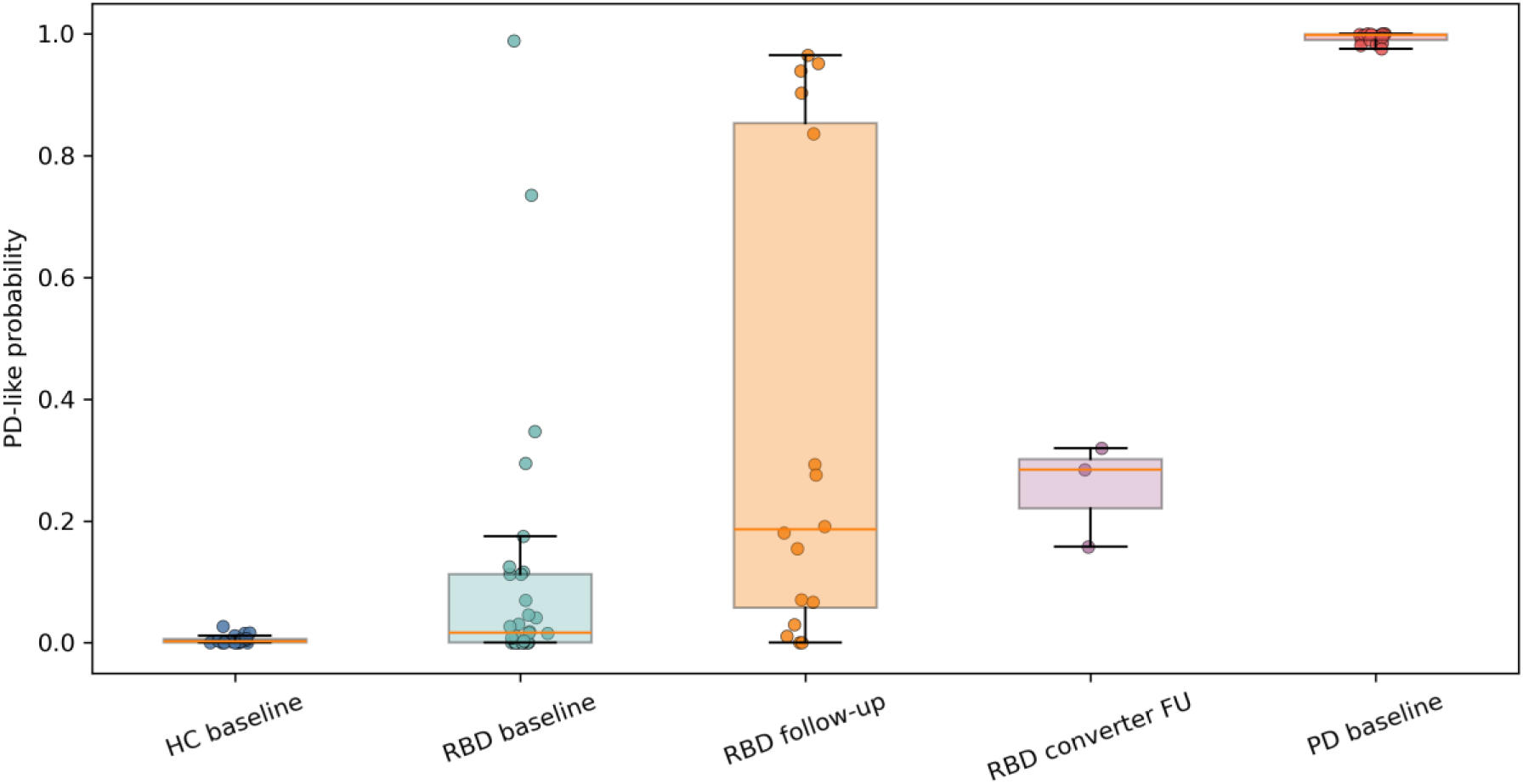
Exploratory PD-like blood transcriptomic projection across baseline and follow-up groups. The score visualizes sample position along an internally learned HC-PD expression axis.

The projection analysis provided a complementary view of iRBD heterogeneity (Figure 5). HC and PD baseline samples occupied opposite ends of the internally learned HC-to-PD axis, whereas baseline iRBD samples were distributed mainly toward the HC-like range but with several higher-scoring individuals. Follow-up iRBD samples showed higher mean and median PD-like probabilities than baseline iRBD, suggesting that some longitudinal samples moved toward a more PD-like blood expression state. However, this pattern was not specific to converter follow-up samples, and the three converters did not represent the highest-scoring projected observations. The projection score should therefore be interpreted as an exploratory molecular axis for visualizing heterogeneity and temporal movement, not as a calibrated risk score or validated predictor of phenoconversion.

**Figure 5.**
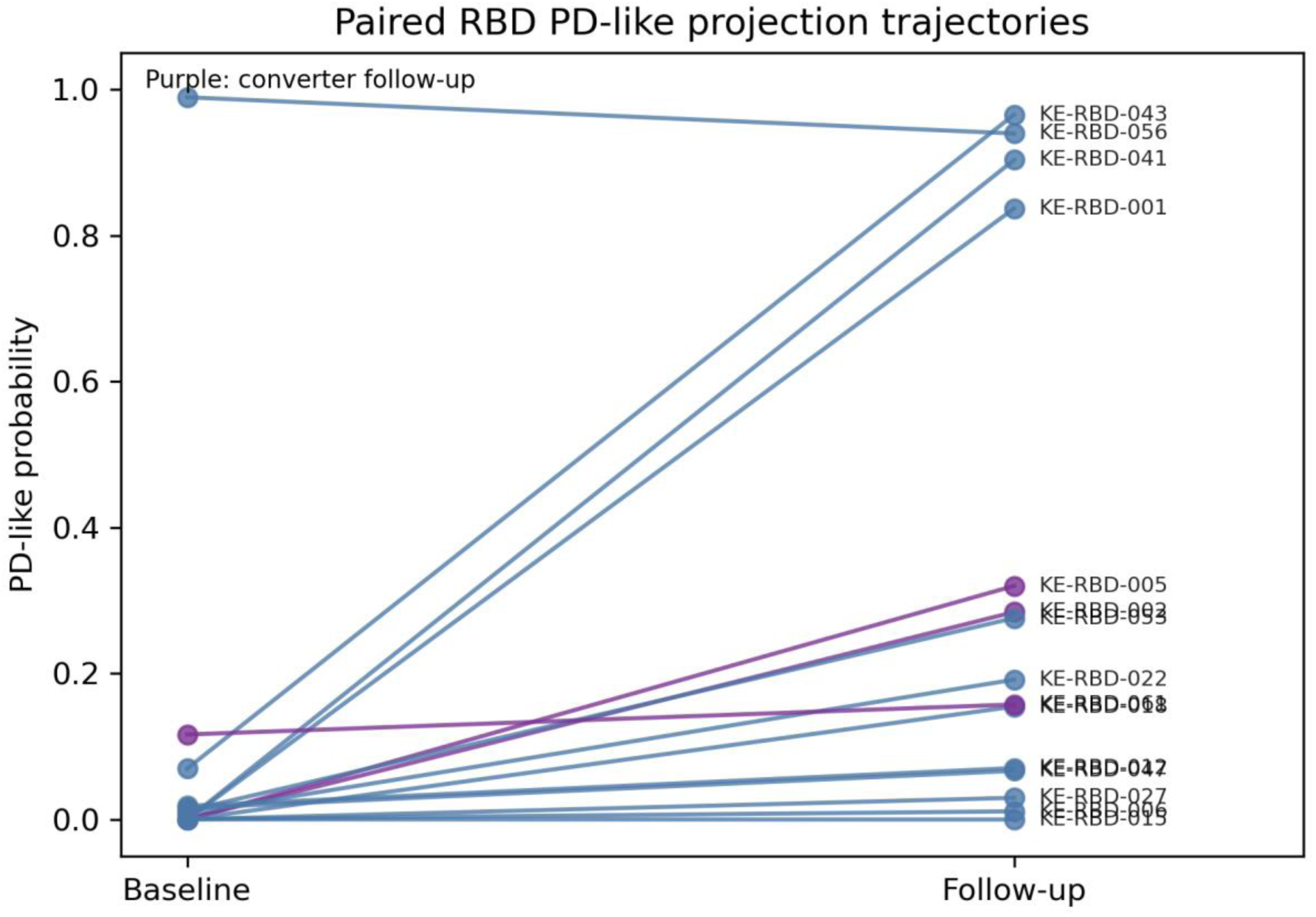
Longitudinal iRBD trajectories on the PD-like projection axis.

### Integrated DESeq2 and ML prioritization identifies candidate PD and exploratory iRBD biomarker panels

Integrated prioritization retained a 24-gene ML-supported PD candidate panel comprising *EGR1, EMILIN2, STAB1, VCAN, ULK1, CCR2, IRS2, SOCS3, CD14, CPVL, MIDN, NFAM1, CREB5, CBX6, NBEAL2, RAB43, PFKFB3, MTMR3, HLA-DRB1, PNPLA2, CSF3R, PLAUR, SLC43A2*, and *CIC*. Heatmap visualization showed a coordinated increase in the standardized expression of many panel genes in PD compared with HC and iRBD, although interindividual variability remained within each diagnostic group. This expression pattern was consistent with the stronger PD discrimination observed in the differential-expression and machine-learning analyses. EGR1 and EMILIN2 showed the highest ML feature-selection stability, followed by STAB1 and VCAN. Consistent with these findings, many panel genes exhibited higher standardized expression in PD than in HC and iRBD, although residual interindividual variability remained within all three groups (Figure 6).

**Figure 6.**
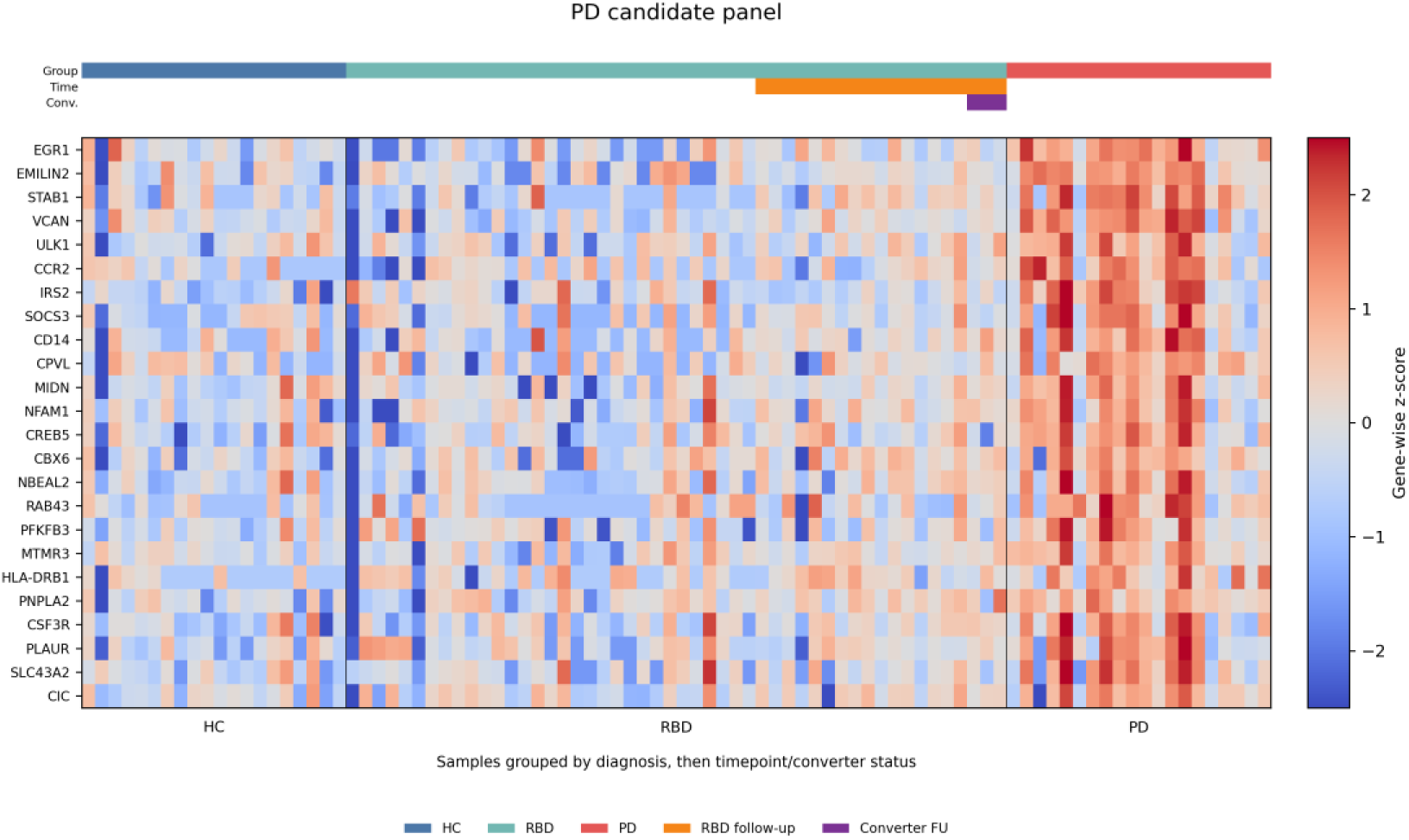
Prioritized PD candidate blood biomarker panel.

The corresponding ML-supported iRBD candidate panel comprised *PHYKPL, DCPS, PELATON, ZC3HC1, BRI3, FEN1, CYSLTR2, TTL, ZBED6, SRGAP2C, CCDC9B, ZNF366, SIGLEC7, ANKRD50, CYP1B1, ZNF839, HMGCR, PALMD, CCDC142, DDX20, LTF, MCM9, MTG1*, and *SETD7*. The heatmap showed a less uniform expression pattern than the PD panel, with many genes displaying lower standardized expression in iRBD than in HC or PD and substantial within-group heterogeneity (Figure 7). This pattern accompanied the single FDR-significant result in the iRBD-versus-HC comparison and the best internal ROC-AUC of 0.584 for HC-versus-iRBD classification.

**Figure 7.**
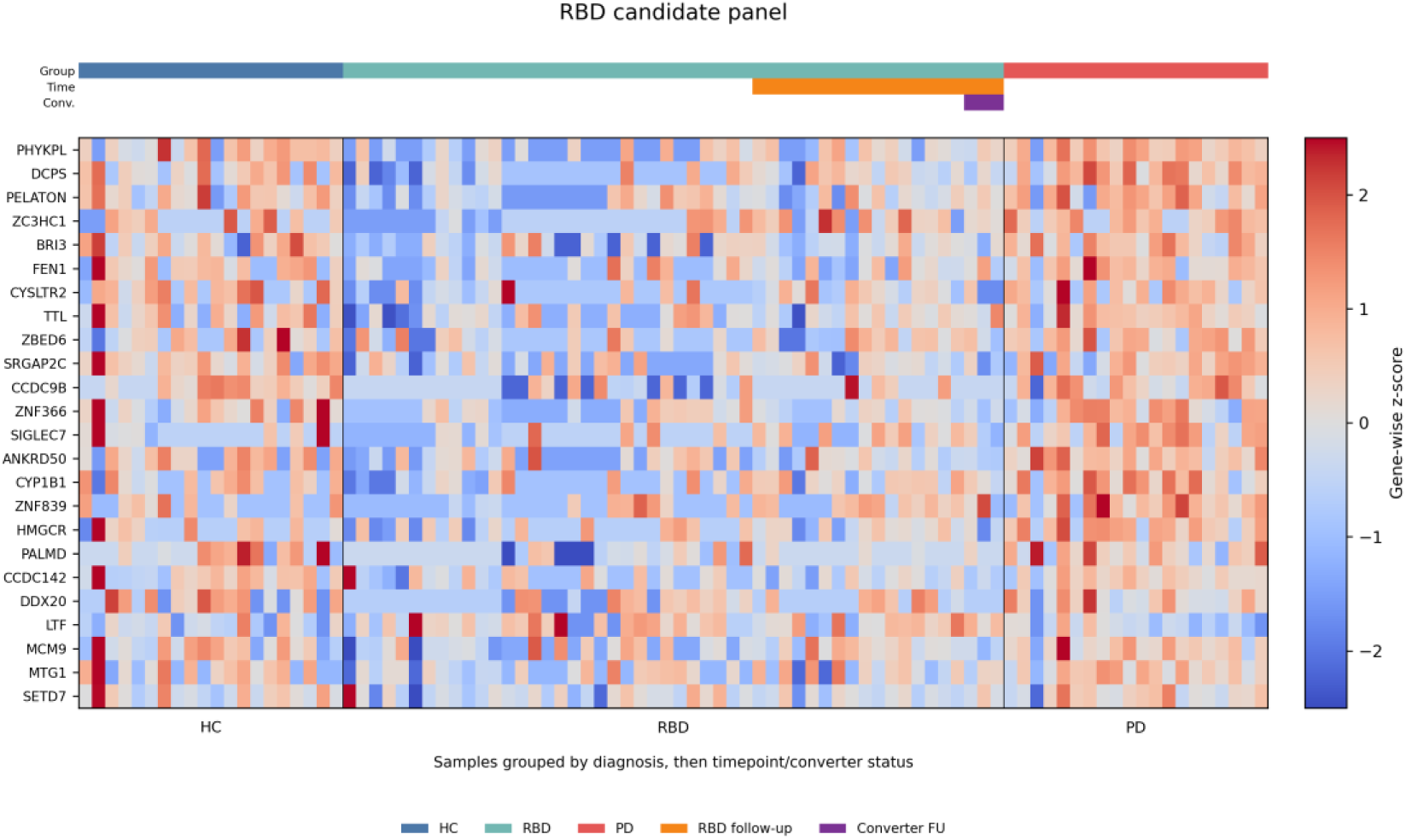
Exploratory iRBD candidate blood biomarker panel.

### Biological annotation of prioritized biomarker candidates

The 24-gene PD candidate panel contained genes associated with several complementary peripheral expression-state processes. Immune, myeloid, hematopoietic, and extracellular-matrix candidates included *VCAN, STAB1, CCR2, CD14, SOCS3, CSF3R, PLAUR, HLA-DRB1, CPVL, NFAM1, NBEAL2*, and *EMILIN2*. Stress-response and transcriptional-regulatory candidates included *EGR1, CREB5, CBX6*, and *CIC*, whereas *ULK1*, *MTMR3*, *PFKFB3*, *IRS2*, *RAB43*, *PNPLA2*, *SLC43A2*, and *MIDN* were related to autophagy, vesicle biology, cellular metabolism, or intracellular signaling.

Over-representation analysis provided pathway-level support for several of these themes. Five KEGG pathways reached FDR significance (Figure 8). The strongest enrichment was observed for the AMPK signaling pathway (four genes: *ULK1, IRS2, CREB5,* and *PFKFB3*; adjusted *P* = 0.0177). Additional significant pathways included longevity-regulating pathway, hematopoietic cell lineage, growth hormone synthesis, secretion and action, and insulin resistance (adjusted *P* = 0.0431 for each). Reactome analysis identified significant enrichment of signaling by leptin, driven by *IRS2* and *SOCS3* (adjusted *P* = 0.0416). The highest-ranked GO Biological Process terms included regulation of hormone levels, cytokine-mediated signaling, cell–cell adhesion, organic-acid transport, and lipid catabolism, although none remained significant after multiple-testing correction.

**Figure 8.**
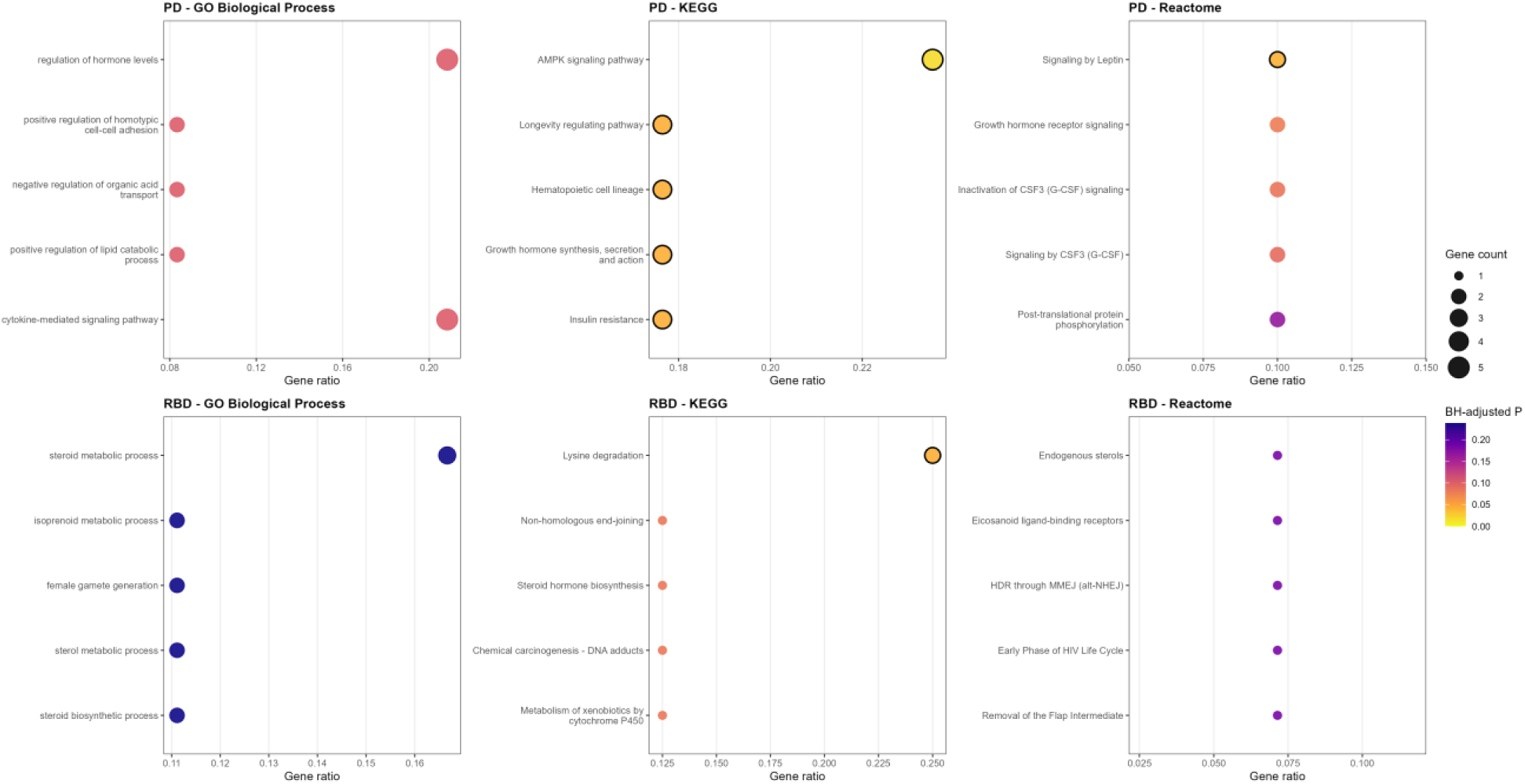
Functional enrichment of the PD and iRBD candidate biomarker panels. Dot plots show the five highest-ranked GO Biological Process, KEGG, and Reactome terms for each 24-gene panel. Dot position represents the gene ratio, dot size represents the number of overlapping panel genes, and color represents the Benjamini–Hochberg-adjusted P value. Black outlines denote terms significant at FDR < 0.05. The enrichment universe comprised 13,925 baseline-expressed genes with Entrez annotations.

The 24-gene iRBD panel represented a more heterogeneous combination of biological processes. Candidates included genes associated with lipid, sterol, and eicosanoid biology (*CYP1B1, HMGCR,* and *CYSLTR2*), DNA maintenance and chromatin regulation (*FEN1, MCM9, SETD7, DDX20,* and *ZC3HC1*), and innate immune or granulocyte-associated biology (*SIGLEC7, LTF,* and *PELATON*). GO Biological Process and Reactome analyses yielded no FDR-significant terms. KEGG analysis identified lysine degradation as the only FDR-significant pathway, driven by *PHYKPL* and *SETD7* (adjusted *P* = 0.0434). Given that this enrichment was based on only two genes, it should be interpreted cautiously.

## Discussion

This study identified a pronounced and internally consistent peripheral blood transcriptional signature of manifest PD, whereas iRBD-associated changes were substantially weaker and more heterogeneous. Differential-expression analysis identified 170 FDR-significant genes in PD versus HC and 85 in PD versus iRBD, compared with only one in iRBD versus HC. This contrast was reflected in the machine-learning results. The best HC-versus-PD model achieved a ROC-AUC of 0.883, and the best iRBD-versus-PD model achieved a ROC-AUC of 0.889, whereas HC-versus-iRBD discrimination remained weak, with a ROC-AUC of 0.584. The agreement between differential expression, out-of-fold classification, candidate-panel visualization, and functional enrichment indicates that the dominant signal in this cohort was associated with clinically manifest PD rather than with iRBD.

The threshold-dependent results further illustrate this distinction. The HC-versus-PD random-forest model correctly classified 17 of 20 HC and 16 of 20 PD samples. For iRBD versus PD, logistic regression correctly classified 21 of 31 iRBD and 17 of 20 PD samples. In contrast, the HC-versus-iRBD model correctly identified only 15 of 31 iRBD samples, and its permutation result did not support discrimination beyond chance. Although the PD-related ROC-AUC estimates were encouraging, their confidence intervals remained relatively wide because of the limited sample size. These estimates should therefore be interpreted as internal evidence of separability rather than as estimates of performance expected in an independent clinical population.

### PD-associated transcriptomic signature and biological interpretation

The 24-gene PD candidate panel captured several biologically related peripheral expression programs. *VCAN*, *STAB1*, *CCR2*, *CD14*, *SOCS3*, *CSF3R*, *PLAUR*, *HLA*-*DRB1*, *CPVL*, *NFAM1*, *NBEAL2*, and *EMILIN2* are associated with myeloid, hematopoietic, immune-signaling, or extracellular-matrix biology. Previous studies have identified transcriptional and functional dysregulation of circulating monocytes in PD, including activation of the CCR2–CCL2 axis and altered expression of inflammatory response genes such as *EGR1* (13,17). Adaptive immune responses to α-synuclein have also been detected in preclinical and early PD, providing additional evidence that peripheral immune activity changes across the disease course (18). Recent experimental work further suggests that immune cells may actively participate in the propagation of synucleinopathy, with intestinal muscularis macrophages modulating α-syn pathology and promoting disease-associated T-cell responses along the gut–brain axis (19). The present panel is therefore consistent with an altered peripheral immune state, although bulk blood RNA-seq cannot distinguish transcriptional activation within individual cells from changes in circulating cell-type proportions.

A second component of the PD panel involved stress responses, transcriptional regulation, autophagy, vesicle biology, and metabolism. Genes *EGR1*, *CREB5*, *CBX6*, and *CIC* represent transcriptional-regulatory candidates, whereas *ULK1*, *MTMR3*, *PFKFB3*, *IRS2*, *RAB43*, *PNPLA2*, *SLC43A2*, and *MIDN* are related to autophagy, intracellular trafficking, nutrient sensing, or cellular metabolism. Autophagic and endolysosomal dysfunction are closely linked to α-synuclein homeostasis and neurodegenerative disease (20). However, the current measurements were obtained from peripheral blood and should not be interpreted as direct evidence of neuronal autophagic dysfunction. They more plausibly reflect systemic metabolic and stress-response programs associated with manifest disease.

Functional enrichment supported this immunometabolic interpretation. The PD panel was significantly enriched for the KEGG AMPK signaling pathway, driven by *ULK1*, *IRS2*, *CREB5*, and *PFKFB3*. Additional significant KEGG terms included hematopoietic cell lineage, longevity regulation, growth-hormone signaling, and insulin resistance. Reactome analysis identified significant enrichment of leptin signaling through *IRS2* and *SOCS3*. The highest-ranked GO terms included cytokine-mediated signaling, regulation of hormone levels, cell adhesion, organic-acid transport, and lipid catabolism, although no GO term remained significant after multiple-testing correction. These findings suggest convergence on hematopoietic and metabolic signaling rather than on a single PD-specific pathway. Because over-representation analysis was based on only 24 selected genes and several pathways were driven by two to four genes, the individual pathway labels should be considered supportive rather than definitive mechanistic evidence.

Comparison with genetic studies further emphasizes the distinction between disease risk and peripheral expression state. Large PD genome-wide association studies have implicated loci including *SNCA*, *GBA1, LRRK2, MAPT, TMEM175, SCARB2*, and the MHC/HLA region (21). At the gene-symbol level, *HLA*-*DRB1* represented the clearest connection between the PD expression panel and a genetically implicated immune region. The remaining panel genes were not prominent canonical PD risk loci. This limited overlap is not unexpected because inherited susceptibility variants and blood RNA expression measure different biological layers. Genetic variants represent lifelong risk, whereas blood transcriptional profiles may capture active immune, metabolic, treatment-related, or disease-state processes. Recent studies have likewise shown that blood transcriptional patterns can reflect clinically relevant PD biology without directly reproducing the genetic architecture of the disease (12,22).

### Interpretation of the iRBD transcriptomic findings

The iRBD findings require greater caution. The 24-gene iRBD panel contained candidates associated with lipid and sterol metabolism, DNA maintenance, chromatin regulation, and innate immune biology, but its heatmap pattern was heterogeneous and frequently reflected lower expression in iRBD relative to HC or PD rather than a uniform iRBD-specific state. GO and Reactome analyses yielded no FDR-significant terms. Lysine degradation was the only significant KEGG term and was driven by only *PHYKPL* and *SETD7*, making this enrichment particularly sensitive to the small panel size. Moreover, none of the iRBD panel genes directly overlapped the principal loci reported by the largest iRBD genome-wide association study, which identified signals near *SNCA*, *GBA*, *TMEM175*, *INPP5F*, and *SCARB2* (23). The iRBD panel should consequently be treated as hypothesis-generating rather than as a validated diagnostic signature.

The weak and heterogeneous iRBD signal should not necessarily be interpreted as evidence against the presence of prodromal molecular alterations. Rather, it may reflect the biological heterogeneity of both prodromal and clinically manifest synucleinopathies. Supported by accumulating clinical, neuroimaging, and histopathological evidence, contemporary models increasingly conceptualize PD as comprising multiple disease trajectories rather than a single uniform pathogenic process. Within the proposed body-first/brain-first framework, premotor iRBD is thought to preferentially identify a body-first trajectory characterized by early involvement of the peripheral autonomic and enteric nervous systems, followed by ascending propagation of α-syn pathology toward the brainstem and higher cortical structures. In contrast, a putative brain-first trajectory may initially involve central or olfactory-associated structures, with RBD developing later during disease progression or remaining absent altogether (3–5).

This distinction has important implications for transcriptomic studies. Although the vast majority of individuals with iRBD ultimately develop a clinically manifest α-synucleinopathy, iRBD is unlikely to represent a universal prodromal stage shared by all future PD patients. Instead, it may preferentially enrich for specific biological trajectories within the broader synucleinopathy spectrum. The PD cohort analysed in the present study was not stratified according to premotor iRBD, autonomic involvement, or proposed body-first and brain-first phenotypes. Consequently, the PD-associated transcriptional signature identified here likely represents a composite signal derived from multiple biological trajectories. If iRBD shares molecular features predominantly with body-first PD, comparison against an unstratified PD cohort could dilute subtype-specific transcriptional patterns and reduce apparent similarity between iRBD and PD.

From this perspective, the broad distribution of iRBD samples along the PD-like projection axis may be biologically informative rather than merely reflecting analytical noise. Scores were lowest in HC, highest in PD, and heterogeneous among baseline iRBD participants, who spanned a continuum from HC-like to more PD-like molecular states. One plausible explanation for the broad distribution of iRBD samples along the PD-like axis is that, in addition to potential subtype-specific biological differences, this projection captures differences in prodromal disease stage. Individuals with iRBD may vary substantially in overall burden of non-motor symptoms, probability of prodromal PD based on the MDS criteria (24), dopaminergic imaging abnormalities, and proximity to phenoconversion (as demonstrated also within our baseline PDBIOM cohort) (25), potentially resulting in a continuum of molecular states between healthy controls and manifest PD. Interestingly, follow-up iRBD observations generally showed higher projection scores than baseline samples, consistent with progressive biological changes occurring during the prodromal phase. This observation is biologically plausible, as longitudinal studies of iRBD typically demonstrate increasing burden of prodromal markers, rising probabilities of prodromal PD, progressive dopaminergic deficits on functional imaging, and ultimately phenoconversion in a subset of individuals (4,26). Although the number of available converters was too small for prognostic conclusions, the observed shift toward more PD-like transcriptional profiles is consistent with this broader pattern of disease progression.

At the same time, the PD-like projection should be regarded as an exploratory framework for visualizing molecular heterogeneity rather than a validated diagnostic or prognostic tool. The projection was derived and evaluated within the same cohort, was not calibrated against time to phenoconversion, and the number of available conversion events was limited. Consequently, projection scores should not be interpreted as individual risk estimates or diagnostic probabilities. Prospective converter-enriched cohorts with repeated sampling, α-syn biomarker assessment, detailed clinical phenotyping, and time-to-event analyses will be required to determine whether longitudinal transcriptomic changes anticipate clinically meaningful progression and whether subtype-specific molecular signatures can be identified before phenoconversion.

### Strengths and limitations

Strengths of the study include the separation of independent baseline samples from longitudinal observations, exclusion of follow-up and post-conversion samples from baseline model development, sequencing-run-aware differential-expression analysis, feature selection and scaling within cross-validation, use of out-of-fold predictions, and integration of differential-expression evidence with ML feature stability. Important limitations include the small cohort, broad confidence intervals, internal validation only, limited permutation resolution, absence of an external replication cohort, incomplete adjustment for blood-cell composition, and possible confounding by age, sex, medication, disease duration, and inflammatory comorbidities. Diagnosis and sequencing run were also incompletely balanced, and the group-informed correction used for exploratory ML and visualization cannot fully resolve this dependence. Furthermore, candidate selection and visualization used the same cohort, meaning that ML support represents internal selection stability rather than independent biomarker validation. These limitations are consistent with current recommendations emphasizing transparent reporting, external validation, and formal risk-of-bias assessment for clinical prediction models (27,28).

Future studies should validate the PD panel using targeted qPCR or digital PCR, evaluate corresponding protein markers where biologically relevant, and replicate performance in independently recruited cohorts. Complete blood counts, cell-type deconvolution, and ideally single-cell profiling will be required to distinguish altered leukocyte composition from cell-intrinsic transcriptional regulation. Also, to address PD heterogeneity it will be necessary to stratify participants according to RBD status, timing of RBD onset, autonomic involvement, and proposed body-first/brain-first features, while enriching longitudinal iRBD cohorts for phenoconversion events. A locked candidate panel should then be evaluated prospectively without feature reselection.

## Conclusion

This study identifies an internally consistent peripheral blood transcriptional signature of manifest PD and prioritizes a 24-gene candidate panel enriched for immunometabolic and hematopoietic processes. While a clear cross-sectional transcriptional signature was readily detectable in manifest PD, the molecular signal observed in iRBD was more variable and did not conform to a single uniform prodromal expression state. The broad distribution of iRBD samples across the PD-like transcriptional axis suggests that molecular differences may already emerge during the prodromal phase, potentially reflecting differences in disease stage, proximity to phenoconversion, and underlying biological subtype. Given that iRBD likely represents a specific prodromal synucleinopathy phenotype rather than the full spectrum of future PD, direct comparisons with unstratified manifest PD cohorts may only partially capture relevant subtype-specific biology. Together, these findings support further validation of peripheral blood biomarkers in PD while highlighting the importance of longitudinal follow-up and biological stratification in future studies of prodromal synucleinopathies.

## Methods

### Study design

This was a peripheral blood RNA-seq biomarker-discovery analysis comparing HC, iRBD, and PD. The analysis distinguished baseline diagnostic biomarker discovery from exploratory longitudinal/projection analyses. The primary biomarker question required independent baseline samples for unbiased cross-sectional discovery. The harmonized metadata were divided into cross-sectional and longitudinal analysis sets before statistical modelling. The primary biomarker-discovery set comprised one independent baseline observation per participant and included 71 samples: 20 HC, 31 iRBD, and 20 PD. This baseline set was used for differential-expression analysis, machine-learning model development, and candidate biomarker-panel prioritization.

Follow-up observations were excluded from baseline differential-expression and machine-learning analyses to prevent repeated measurements from the same participant from being treated as statistically independent observations. Nineteen iRBD follow-up observations, including three observations collected after phenoconversion, were retained exclusively for exploratory longitudinal and PD-like projection analyses. Because of the limited number of converter observations, these analyses were considered descriptive and hypothesis-generating and were not used to estimate diagnostic performance or prospective conversion risk.

### Study cohorts and participant selection

Individuals with PD, iRBD and HC were recruited from three ongoing longitudinal cohorts established at the Department of Neurology, P. J. Safarik University and L. Pasteur University Hospital, Kosice, Slovakia. All participants provided written informed consent prior to enrolment. The studies were approved by the local ethics committee and conducted in accordance with the Declaration of Helsinki.

20 patients with PD were recruited at the local Movement Disorders Centre within the framework of the Central European Group on Genetics of Movement Disorders (CEGEMOD) consortium, a multicentre registry of movement disorder patients and controls established through a collaborative network of tertiary movement disorder centres across Central Europe (29). PD diagnosis was established by movement disorder specialists according to the Movement Disorder Society clinical diagnostic criteria (30).

31 individuals with iRBD were recruited from the ongoing Parkinson’s Disease BIOMarker (PDBIOM) cohort between 2018 and 2026 (25). Recruitment was performed through a media campaign focused on RBD symptoms and their association with neurodegenerative diseases. Potential participants underwent a two-step screening procedure consisting of the REM Sleep Behaviour Disorder Single-Question Screening (RBD1Q) followed by the REM Sleep Behaviour Disorder Screening Questionnaire (RBDSQ). Subjects scoring ≥5 points on the RBDSQ subsequently underwent overnight video-polysomnography (v-PSG). The diagnosis of iRBD was confirmed according to the American Academy of Sleep Medicine (AASM) criteria (31), supplemented by electromyographic assessment following the Sleep Innsbruck Barcelona Group (SINBAR) recommendations (32). Individuals with manifest neurodegenerative disease at baseline were excluded. Since 2020, participants have been prospectively followed, with follow-up assessments performed approximately 2–5 years after baseline, including evaluation of phenoconversion to clinically manifest neurodegenerative disease. 19 follow-up samples were available for the present study, including three individuals who phenoconverted to PD.

20 healthy controls were recruited from both the PDBIOM and PARCAS cohorts. PDBIOM controls underwent the same screening protocol as iRBD participants but had negative v-PSG examinations and did not fulfil diagnostic criteria for RBD. Additional controls were recruited from the PARkinson’s disease associated Colonic Alpha-Synuclein biomarker study (PARCAS) cohort, a prospective longitudinal study established between 2014 and 2018 among individuals undergoing diagnostic colonoscopy. Participants were enrolled based on the presence of gastrointestinal symptoms or other potential prodromal features of PD (33) and were followed prospectively between 2018 and 2024 (34). Both the PDBIOM and PARCAS cohorts underwent comprehensive evaluation of risk factors and prodromal markers according to the Movement Disorder Society (MDS) research criteria for prodromal PD (24). For the present study, HC were selected from individuals who did not fulfil criteria for prodromal PD and whose estimated prodromal PD probability remained comparable to the age-related background risk of the general elderly population after comprehensive assessment.

### Sample collection and quality assessment

Peripheral venous blood samples in EDTA tubes were collected, and total RNA was isolated from each participant and used for downstream RNA sequencing and transcriptomic analysis aimed at identifying early molecular biomarkers predictive of Parkinson’s disease development.

The quality of total RNA was assessed using the Agilent RNA 6000 Pico Kit on the Agilent 2100 Bioanalyzer (Agilent Technologies, Santa Clara, USA) according to the internal standard operating procedure (SOP-SK-5-2). The RNA Integrity Number (RIN) was determined using the Agilent 2100 Expert software (version B.02.10.SI764). Although several RNA samples displayed partial degradation and lower concentration, all selected samples met the minimum quality requirements for downstream transcriptome analysis.

### RNA-seq preprocessing and gene-level quantification

Raw paired-end RNA-seq reads were processed using a standardized quality-control and quantification workflow. Initial read quality was assessed using FastQC (version 0.12.1), including per-base sequence quality, GC-content distribution, sequence duplication, adapter contamination, and overrepresented sequences.

Adapter removal, quality trimming, and read filtering were performed using fastp (version 1.0.1). Reads failing the predefined quality or ambiguous-base thresholds were excluded. Post-filtering quality metrics were subsequently evaluated to confirm adequate read quality and adapter removal.

Filtered reads were aligned to the human GRCh38 reference genome using STAR (version 2.7.11b) in two-pass mode. Total mapping rate, uniquely mapped reads, multimapping reads, and other alignment metrics were extracted from the STAR log files and used for sample-level quality assessment.

Transcript abundance was estimated from the filtered paired-end reads using Salmon (version 1.10.3) in mapping-based mode with GC-bias correction. Transcript-level abundance estimates were imported into R using tximport and summarized to gene-level estimated counts for downstream differential-expression analysis. Quality-control, filtering, and alignment metrics were aggregated across samples using MultiQC (version 1.34).

### Differential-expression analysis

Baseline differential expression was performed with DESeq2 (version 1.46.0) using design ∼ source dataset + group. Primary contrasts were PD vs HC, PD vs RBD, and RBD vs HC.

Variance-stabilized expression matrices were generated for baseline and combined baseline-plus-follow-up data. Source correction removed source-dataset effects while preserving diagnostic group structure. This group-informed setup is reported as a limitation. Genes were considered significantly differentially expressed if they met an adjusted p-value (padj, FDR) ≤ 0.05.

To ensure biological relevance, the log_2_ fold change (LFC) threshold was set to |LFC| ≥ 1. Only genes meeting both statistical (FDR ≤ 0.05) and fold-change criteria were retained for downstream analysis.

### Machine-learning biomarker analysis

Machine-learning analyses were performed using the source-corrected variance-stabilized expression matrix restricted to independent baseline samples. Follow-up and converter observations were excluded from model training. The baseline ML dataset included 71 samples: 20 healthy controls, 31 isolated RBD patients, and 20 PD patients.

Three binary classification tasks were evaluated: HC vs PD, HC vs RBD, and RBD vs PD. Models were implemented in scikit-learn (version 1.5.1) pipelines to reduce information leakage. Each pipeline included fold-internal univariate feature selection using ANOVA F-statistics (SelectKBest) before model fitting. Logistic-regression models additionally included standard scaling and class-balanced logistic regression using the liblinear solver. Random-forest models used class-balanced random forests. The analysis evaluated feature-selection sizes of 50, 250, and 500 genes, logistic-regression regularization strengths C = 0.01, 0.1, 1 and random forests with 300 trees and either unrestricted depth or maximum depth of 5.

Classifier performance was estimated using nested stratified cross-validation. The outer loop generated out-of-fold predicted probabilities using up to five stratified folds, depending on the smallest class size. Hyperparameters were selected within each outer training fold using an inner stratified grid search optimized for ROC-AUC. Performance was reported as ROC-AUC, precision-recall AUC, balanced accuracy, sensitivity, specificity, F1 score, and confusion matrices. Confidence intervals for ROC-AUC and precision-recall AUC were estimated using 500 bootstrap resamples of the out-of-fold predictions. Exploratory permutation testing was performed with 50 label permutations.

### PD-like projection analysis

To explore whether isolated RBD samples showed evidence of a shift toward a PD-like peripheral blood transcriptional state, we generated an exploratory PD-like projection score. This analysis was designed as a visualization and hypothesis-generating approach rather than as a diagnostic or prognostic model.

A binary HC vs PD classifier was trained using only independent baseline samples from the source-corrected variance-stabilized expression matrix. RBD samples, RBD follow-up samples, and converter follow-up samples were not used for model training. This ensured that the projection axis was defined only by the contrast between healthy controls and manifest PD, and that RBD samples could subsequently be positioned along this axis without directly contributing to the model.

The trained HC vs PD model was then applied to the combined baseline-plus-follow-up expression matrix. For each sample, the model generated a predicted probability of belonging to the PD class. This probability was interpreted as a PD-like transcriptional projection score, with values closer to 0 indicating a more HC-like blood expression profile and values closer to 1 indicating a more PD-like profile. Scores were summarized across HC baseline, RBD baseline, RBD follow-up, RBD converter follow-up, and PD baseline groups.

For longitudinal visualization, RBD patients with available baseline and follow-up observations were connected across time points to assess whether individual trajectories shifted toward or away from the PD-like axis. Converter follow-up samples were retained as a separate annotated subgroup in plots, but the number of converters was too small for formal prognostic modeling. Therefore, converter trajectories were interpreted descriptively only.

### Candidate biomarker-panel prioritization

Candidate biomarker panels were prioritized by integrating differential-expression evidence with ML-derived feature stability. For each binary task, logistic-regression feature stability was assessed across stratified cross-validation folds. Within each fold, the top 250 genes were selected using ANOVA F-statistics before model fitting, and genes were ranked according to the number and proportion of folds in which they were selected.

ML-derived feature stability was then integrated with DESeq2 evidence, concordant direction of effect across relevant contrasts, effect size, statistical significance, and protein-coding/protein-like gene annotation. PD panel prioritization used DESeq2 evidence from PD vs HC and PD vs RBD, together with ML feature stability from HC vs PD and RBD vs PD. RBD panel prioritization used evidence from RBD vs HC and RBD vs PD, together with ML feature stability from HC vs RBD and RBD vs PD.

The final ML-informed biomarker panels were therefore selected as internally prioritized candidate panels rather than externally validated diagnostic signatures. Heatmaps display gene-wise z-scores for the full 24-gene PD and RBD panels across HC, RBD, RBD follow-up, converter follow-up, and PD samples.

### Enrichment analysis

Over-representation analysis was performed separately for the 24-gene PD and RBD candidate panels using clusterProfiler, org.Hs.eg.db, Reactome, and KEGG pathway annotations in R (version 4.5.2). The analysis universe comprised 13,925 Entrez-mapped genes represented in the baseline expression dataset. GO Biological Process, KEGG, and Reactome terms were tested using the hypergeometric framework, with Benjamini–Hochberg correction performed separately within each database. Terms with an adjusted *P* value below 0.05 were considered statistically significant.

## Data Availability

All data produced in the present study are available upon reasonable request to the authors.

## Data availability

Due to the sensitive nature of potentially identifiable protected health information of participants, deidentified clinical data will be made available upon request pursuant to institutional approvals for a Data Use Agreement or equivalent agreement as appropriate.

## Code availability

Code used for data processing and analysis is available from the corresponding authors upon reasonable request.

## Ethics approval

2023/EK/05020 - University Hospital L. Pasteur, Kosice, Slovakia

